## Supplemental Information for "Measuring changes in *Plasmodium falciparum* census population size in response to sequential malaria control interventions"

### SUPPLEMENTAL METHODS

##### *Var* genotyping and sequence analysis

For *var* genotyping, the sequence region within the *var* genes encoding the DBLα domain of PfEMP1 were amplified in a single-step PCR from genomic DNA using primers for multiplexed sequencing as reported in Rask et al. (Rask et al., 2016). We coupled template-specific degenerate primer sequences targeting homology block 2 (forward primer: DBLαAF, 5'-GCACGMAGTTTYGC-3') and homology block 3 (reverse primer: DBLαBR, 5'-GCCCATTCSTCGAACCA- 3') (Bull et al., 2005; Rask et al., 2010) with a GS FLX Titanium primer sequence. Each of the forward and reverse DBLα primers were barcoded with a unique 10 bp multiplex identifier (MID) tag published by (Roche, 2009). For additional details on the validation of these primers for amplification of sequences of the appropriate length (~477 bp) using *P. falciparum* reference strains (3D7, Dd2, and HB3) see Rask et al. (Rask et al., 2016).

Each PCR reaction was prepared in a total volume of 40 µL consisting of 0.5x buffer, 2 mM of $MgCl_2$, 0.07 mM of dNTPs, 0.375 µM of each primer (DBLαAF, DBLαBR), 3 units of GoTaq G2 Flexi DNA polymerase (Promega), and 2 µL of isolate genomic DNA. The PCR cycling conditions involved 95°C for 2 min, followed by 30 cycles of 95°C for 40 seconds, 49°C for 90 seconds, 65°C for 90 seconds, and a final extension step of 65°C for 10 min. Positive controls (laboratory genomic *P. falciparum* DNA) and a negative control (no template) were included for quality assurance. The PCR products were purified using the SPRI method (solid-phase reversible immobilization) (Agencourt, AMPure XP). Purified PCR product concentrations were measured using the Quant-iT PicoGreen dsDNA kit as per the manufacturer's instructions (Invitrogen). We assayed fluorescence intensity using a Perkin-Elmer VICTOR X3 multilabel plate reader, with fluorescein excitation wavelength of ~480 nm and emission of ~520 nm wavelength. Amplicons were then pooled equimolarly with each pool consisting of up to 106 isolates, all with unique MID tags. The libraries were then prepared using the KAPA HiFi HotStart Ready Mix (Kapa Biosystems) and sequenced on an Illumina platform using the MiSeq Reagent Kit v3 (600 cycle; 2×300bp paired-end) (New York University Genome Technology Center, New York, NY, USA; Australian Genome Research Facility, Melbourne, Australia) (Figure supplement 1).

The raw sequence data was then cleaned using our published customized bioinformatic pipeline (https://github.com/UniMelb-Day-Lab/DBLaCleaner) (He et al., 2018). This pipeline was used to de-multiplex and merge the paired-end reads as well as remove low-quality sequences and chimeras using several filtering parameters (see Figure supplement 2 for additional details). These steps resulted in a total of 291,783 cleaned DBLα sequences for the 2,572 *P. falciparum* isolates sequenced (Figure supplement 2, Table supplement 2). To identify the unique DBLα types, we then clustered these cleaned DBLα sequences with 241,693 DBLα sequences available from this Bongo study, as well as the *P. falciparum* reference strains (3D7, Dd2, and HB3) included as positive controls (He et al., 2018; Pilosof et al., 2019; Rorick et al., 2018), at the standard 96% sequence identity (https://github.com/UniMelb-Day-Lab/clusterDBLalpha) (Barry et al., 2007; Day et al., 2017; Ruybal-Pesántez et al., 2017). Our dataset was then further curated by translating the DBLα types (N = 68,503) into amino acid sequences and removing any DBLα types that could not be translated (i.e., contained a stop codon) (N = 135; 0.2%). The remaining DBLα types were then assigned to their most likely DBLα domain class (i.e., DBLα0, DBLα1, or DBLα2) using a hidden Markov model, and further classified based on the association of specific domain classes with semi-conserved upstream promoter sequences (ups) as either upsA or non-upsA DBLα types (https://github.com/UniMelb-Day-Lab/classifyDBLalpha) (Ruybal-Pesántez et al., 2017).

### SUPPLEMENTAL FIGURES

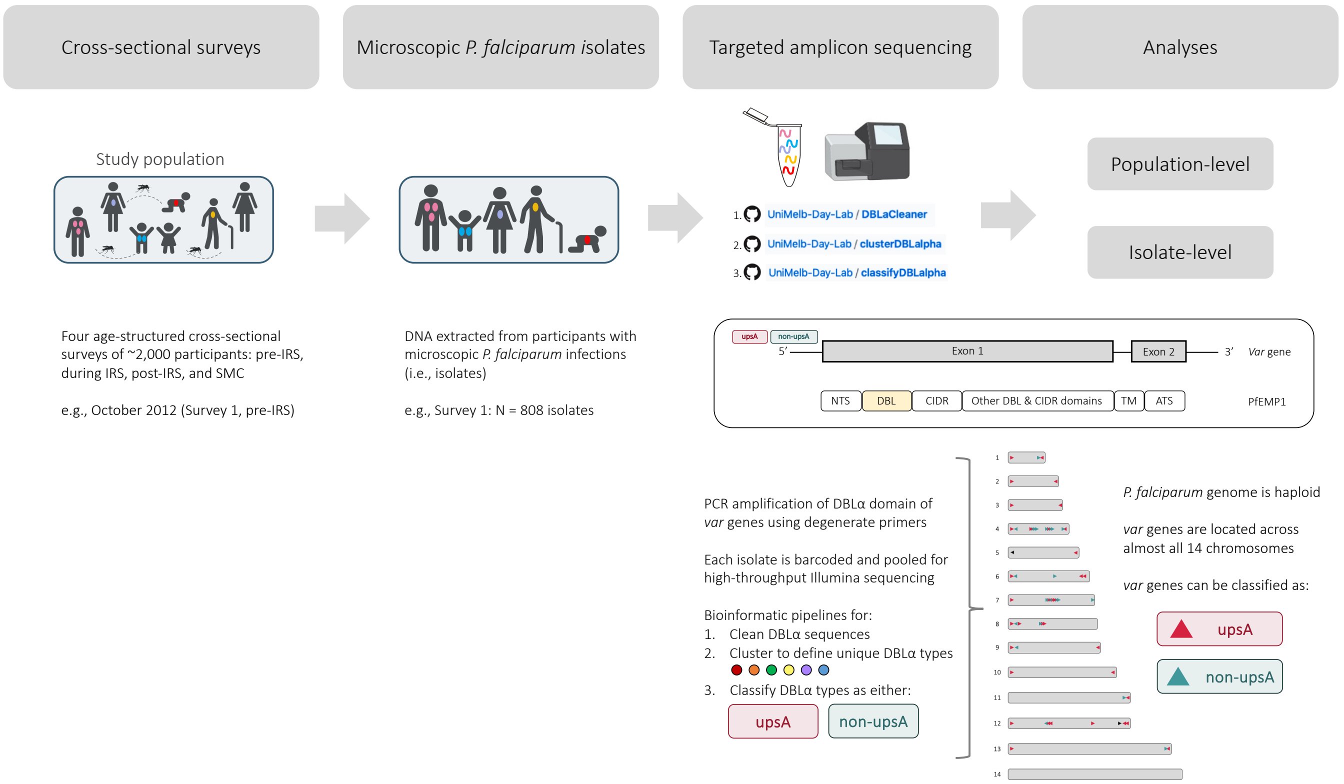


**Figure supplement 1. Schematic diagram of the *var* genotyping (i.e., *var*coding) approach.** For additional details about each step, see Materials and Methods and Supplemental Methods. A schematic insert of the *var* gene locus and PfEMP1 has been included as defined in Rask et al. (Rask et al., 2010), with its N-terminal segment (NTS), Duffy binding-like (DBL) domains, cysteine-rich interdomain regions (CIDR), one transmembrane region (TM), and the acidic terminal segment (ATS). The Illumina MiSeq sequencer stock image was created with BioRender.com.

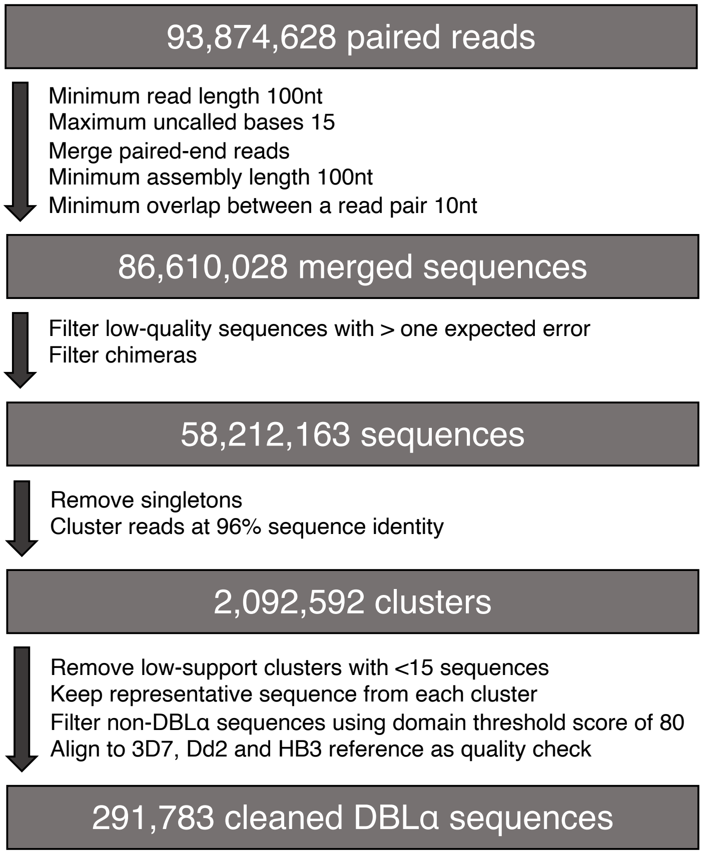


**Figure supplement 2. Bioinformatic sequence data processing flowchart.** The flowchart shows the bioinformatic process to clean the raw de-multiplexed paired reads, with details on the filtering parameters utilized at each step. This customized bioinformatic pipeline is described in detail in He et al. (He et al., 2018).

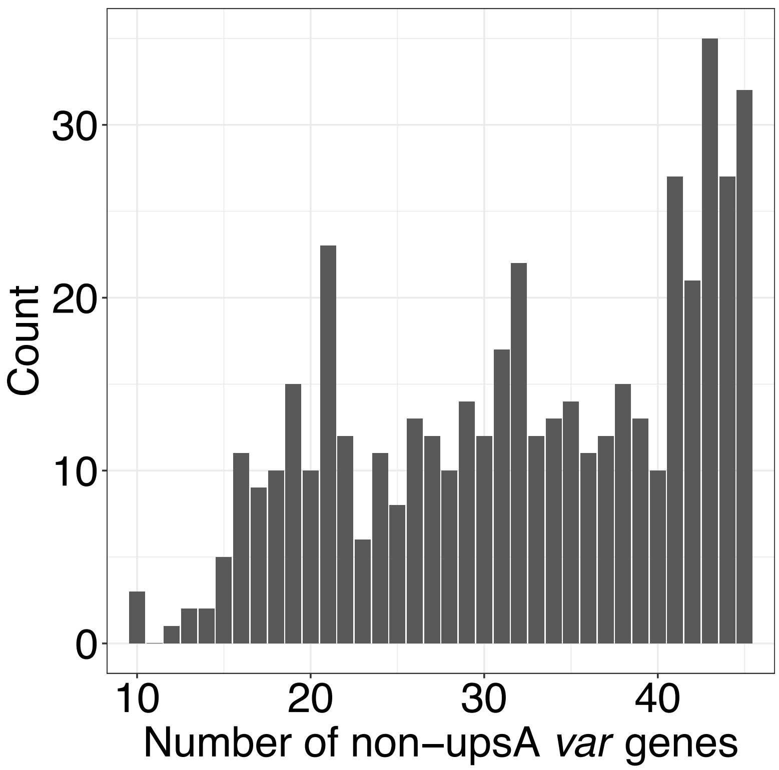


**Figure supplement 3. Histogram of the number of non-upsA DBLα *var* gene types sequenced per repertoire for those isolates with monoclonal infections (MOI = 1)** (Labbé et al., 2023)**.** The molecular sequences used to derive this repertoire size distribution were previously sequenced from isolates sampled during six cross-sectional surveys made from 2012 to 2016 in Bongo District, Ghana (He et al., 2018; Pilosof et al., 2019; Ruybal-Pesántez et al., 2022; Tiedje et al., 2022). These isolates were estimated to be monoclonal infections (i.e., human hosts estimated to be infected by a single *P. falciparum* clone, MOI = 1), based on a cut-off value of 45 non-upsA DBLα types. This cut-off was selected based on the median number of non-upsA DBLα types identified for the 3D7 laboratory isolate included as a control during *var*coding (Ghansah et al., 2023). Note a version of this figure was previously published in Labbé et al. (Labbé et al., 2023)**.**

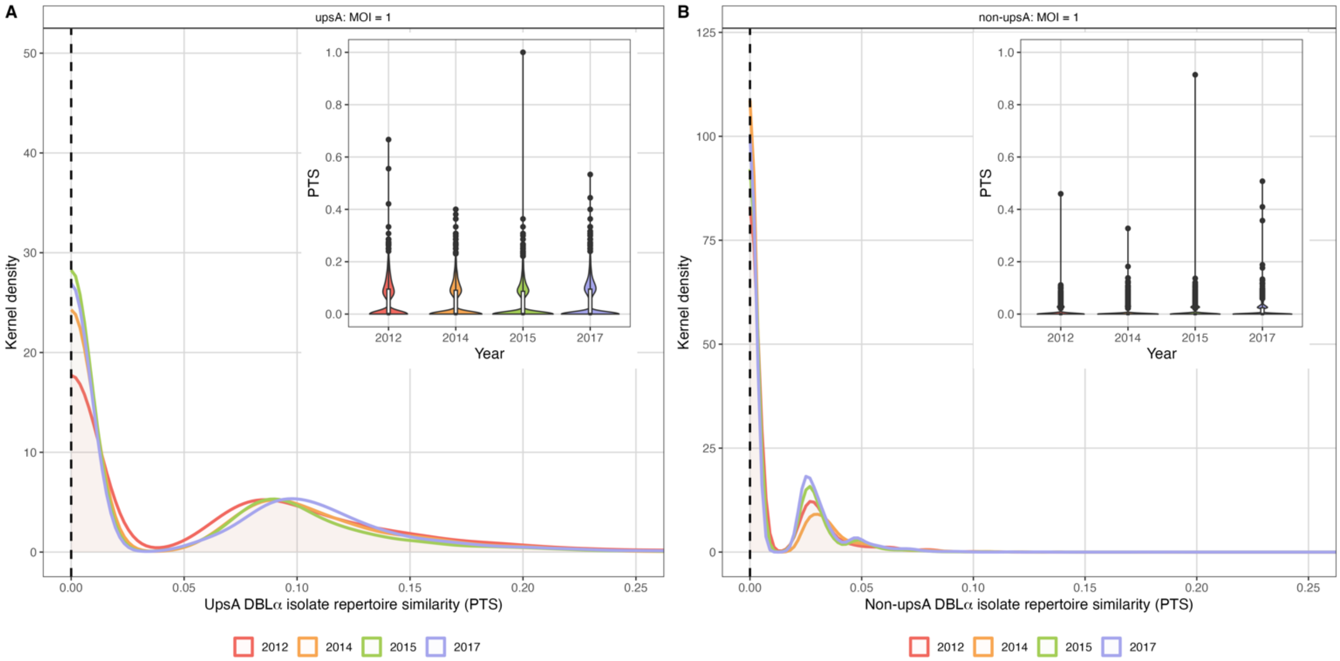


**Figure supplement 4. Sharing of upsA and non-upsA DBLα types among the DBLα isolate repertoires for those isolates with an estimated $MOI_{var}$ equal to one ($MOI_{var}$ = 1) in 2012 (pre-IRS, red), 2014 (during IRS, orange), 2015 (post-IRS, green), and 2017 (SMC, purple).** The overlapping density and violin plots (upper right-hand corners) show the distribution of PTS scores (i.e., DBLα isolate repertoire similarity) between the (**A**) upsA and (**B**) non-upsA DBLα isolate repertoires with an $MOI_{var}$ = 1 in each survey. The PTS scales in the density plots have been zoomed-in to provide better visualization of the PTS distributions for the upsA and non-upsA DBLα types. The black dashed lines in the density plots indicate the median PTS scores for the upsA and non-upsA DBLα types in each survey, which were all equal to zero. *Note:* In the PTS violin plots the central box plots indicate the medians (centre line), interquartile ranges (IQR, upper and lower quartiles), whiskers (1.5x IQR), and outliers (points).

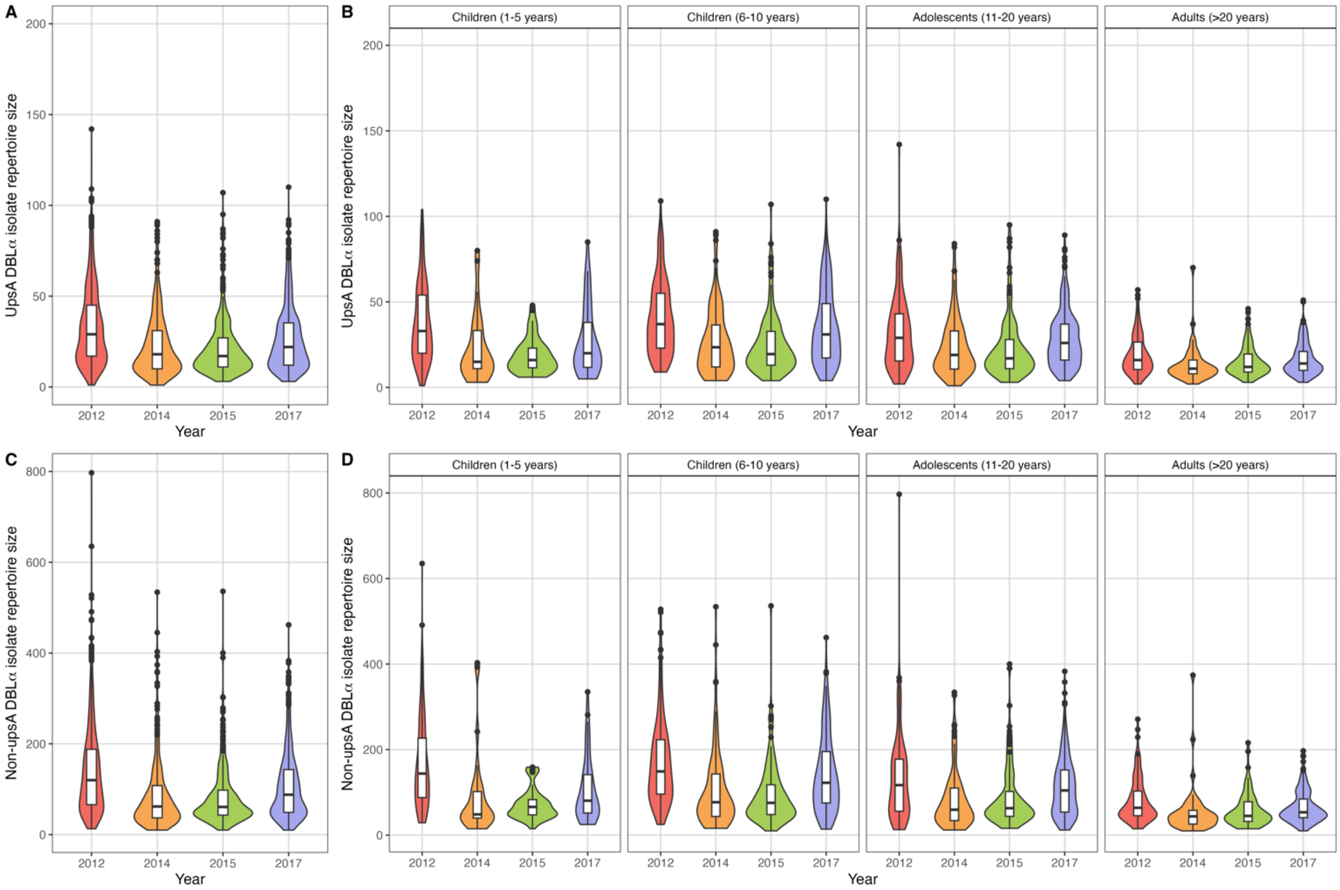


**Figure supplement 5. UpsA and non-upsA DBLα isolate repertoire sizes in 2012 (pre-IRS, red), 2014 (during IRS, orange), 2015 (post-IRS, green), and 2017 (SMC, purple)**. Violin plots showing the frequency distributions of the upsA and non-upsA DBLα isolate repertoire sizes for the (**A, C**) study population and (**B, D**) for all age groups (years) in each survey. The width of each violin plot illustrates the relative frequency of the upsA and non-upsA DBLα repertoire sizes in each survey. *Note:* The y-axis range for the upsA and non-upsA DBLα isolate repertoire sizes are different. The central box plots indicate the median upsA and non-upsA DBLα repertoire sizes (centre line), interquartile ranges (IQR, upper and lower quartiles), whiskers (1.5x IQR), and outliers (points) for each violin plot.

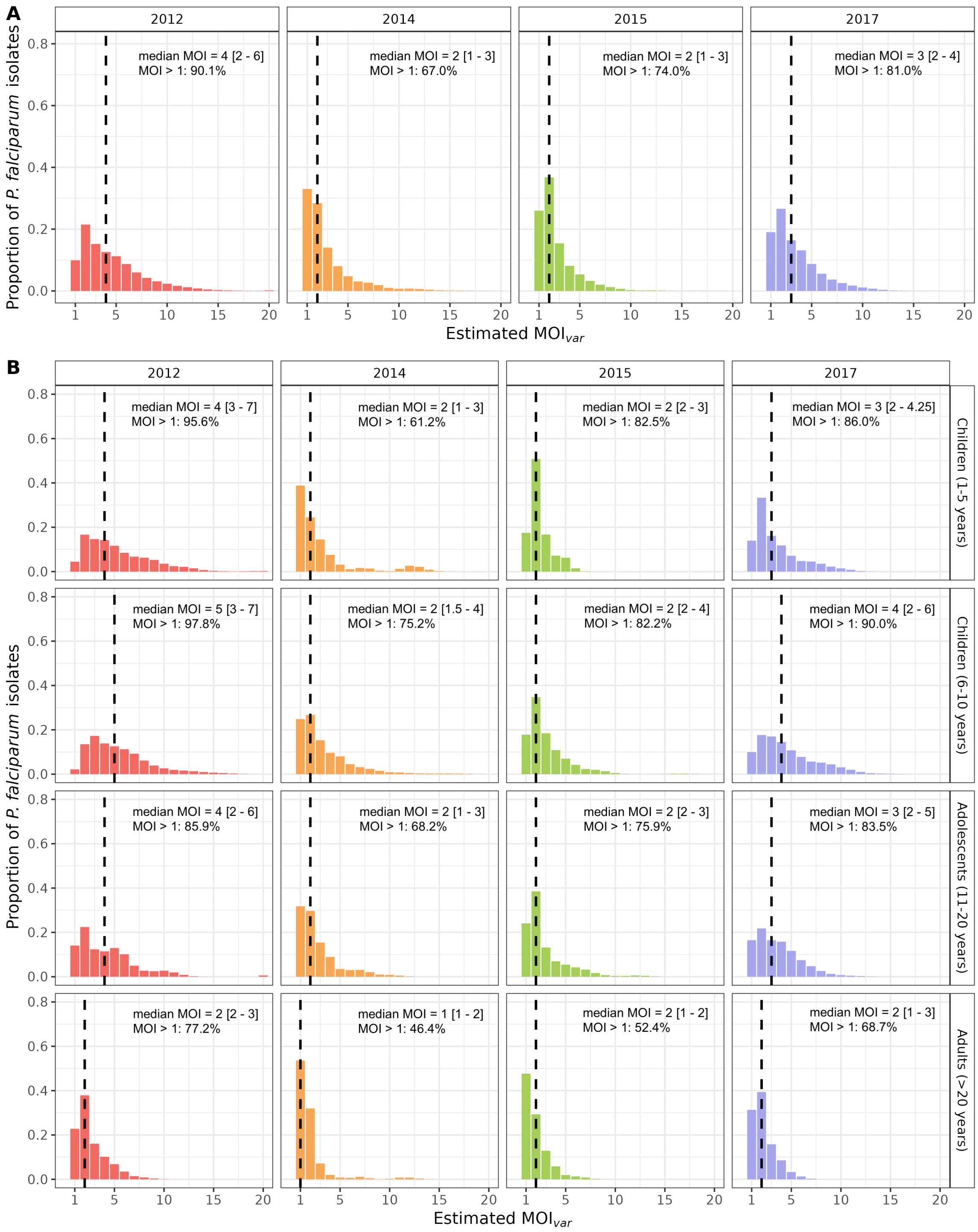


**Figure supplement 6. $MOI_{var}$ distributions in 2012 (pre-IRS, red), 2014 (during IRS, orange), 2015 (post-IRS, green), and 2017 (SMC, purple) based on the mixture distribution approach.** Estimated $MOI_{var}$ distributions for the (**A**) study population and (**B**) for all age groups (years) in each survey for those isolates with DBLα sequencing data (Table supplement 2 and 6). The median $MOI_{var}$ values are indicated with the black dashed lines and have been provided in the top right corner (median $MOI_{var}$ value [interquartile range, upper and lower quartiles]) along with the percentage of *P. falciparum* infections that were multiclonal ($MOI_{var} > 1$) in each survey and age group (years).

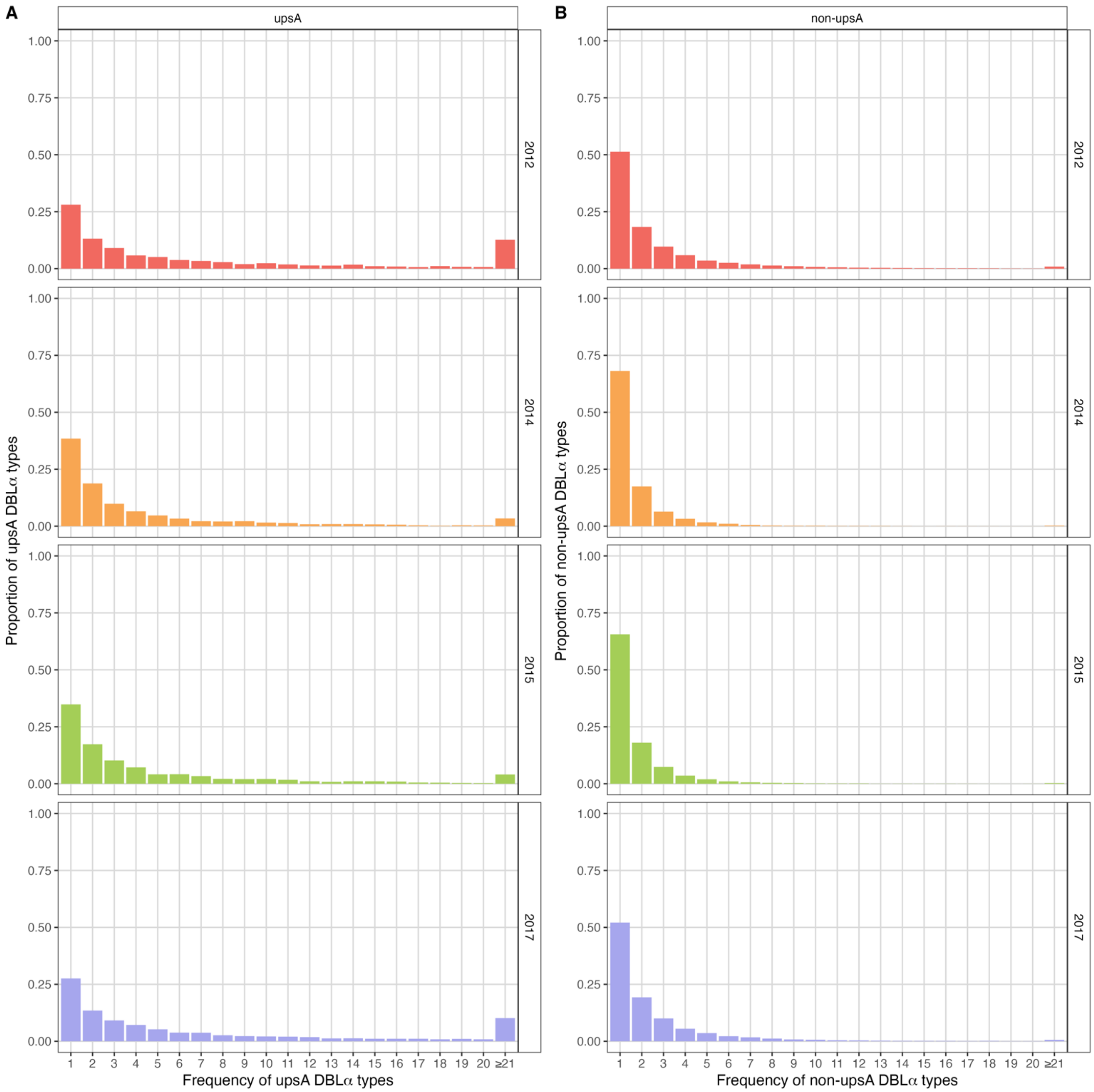


Figure supplement 7. Frequency distributions for the (A) upsA and (B) non-upsA DBLα types in 2012 (pre-IRS, red), 2014 (during IRS, orange), 2015 (post-IRS, green), and 2017 (SMC, purple).

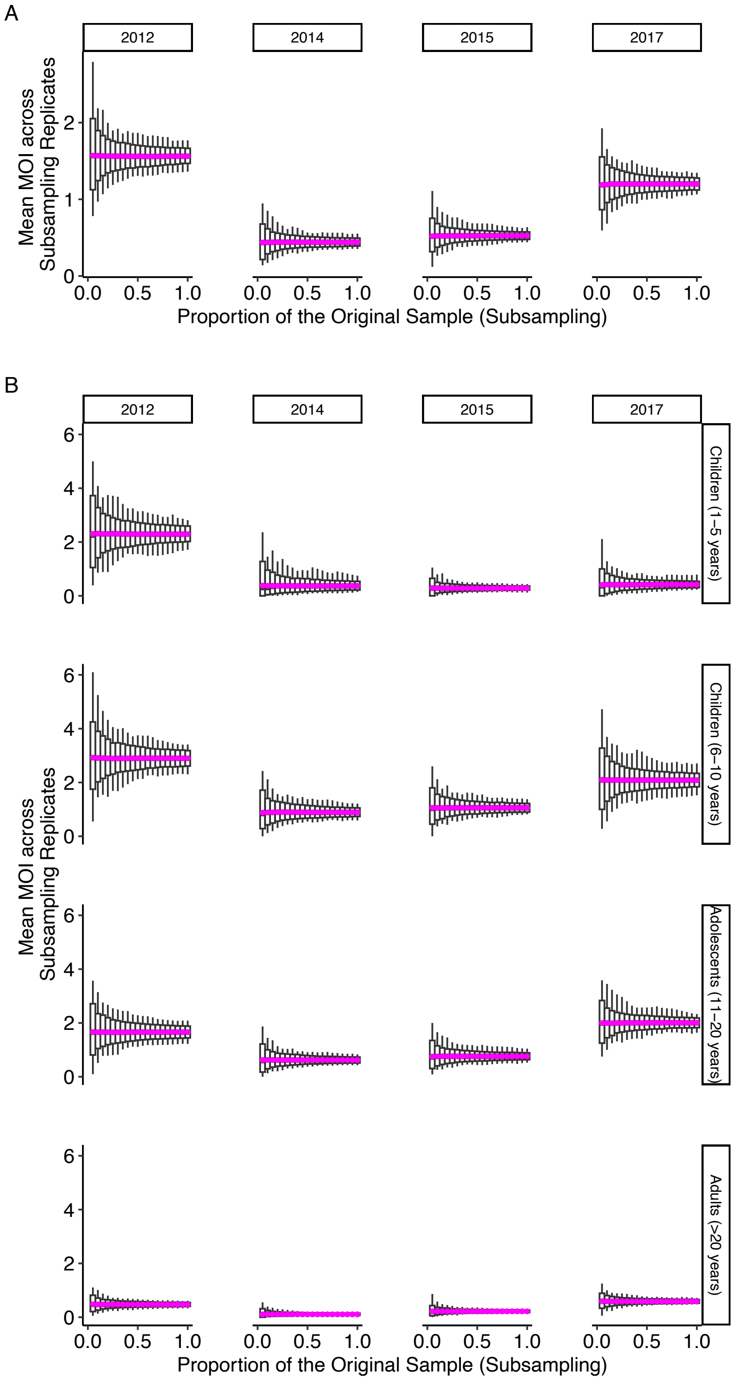


**Figure supplement 8. Mean MOI averaged over all sampled individuals for the subsampling replicates in 2012 (pre-IRS), 2014 (during IRS), 2015 (post-IRS), and 2017 (SMC).** The mean MOI (pink dots) for the (**A**) study population and (**B**) for all age groups (years) in each survey is stable relative to the sampling size or sampling depth, which allows for the extrapolation from the census population size of our population sample to that of the whole population of local hosts. For each sampling depth, we generate 1,000 subsampling replicates with replacement. Minimum, 5% quantile, median, 95% quantile, and maximum values are shown in the boxplot.

### SUPPLEMENTAL TABLES

**Table supplement 1.** Age group (years) breakdown and parasitological characteristics of the participants surveyed in Bongo, Ghana in each survey (i.e., 2012, 2014, 2015, 2017).

| | October 2012 (pre-IRS) | October 2014 (IRS) | October 2015 (post-IRS) | October 2017 (SMC) |
|---|---|---|---|---|
| Number of participants surveyed [a] | 1923 | 1866 | 2022 | 1915 |
| Age groups (years) [b] | | | | |
| Children: 1-5 years | 356 (18.5) | 216 (11.6) | 405 (20.0) | 354 (18.5) |
| Children: 6-10 years | 395 (20.5) | 421 (22.5) | 409 (20.2) | 358 (18.7) |
| Adolescents: 11-20 years | 413 (21.5) | 468 (25.1) | 467 (23.1) | 489 (25.5) |
| Adults: >20 years | 759 (39.5) | 761 (40.8) | 741 (36.7) | 714 (37.3) |
| Microscopic *P. falciparum* prevalence [c] | **808 (42.0)** | **430 (23.0)** | **545 (27.0)** | **789 (41.2)** |
| Children: 1-5 years | 173 (48.6) | 37 (17.1) | 63 (15.6) | 49 (13.8) |
| Children: 6-10 years | 243 (61.5) | 142 (33.7) | 167 (40.8) | 184 (51.4) |
| Adolescents: 11-20 years | 202 (48.9) | 162 (34.6) | 169 (36.2) | 304 (62.2) |
| Adults: > 20 years | 190 (25.0) | 89 (11.7) | 146 (19.7) | 295 (35.3) |
| Microscopic *P. falciparum* density [d] | **520 [160-1,640] (40-126,040)** | **200 [80-600] (40-73,360)** | **320 [120-1,800] (40-113,520)** | **440 [160-1,720] (40-97,520)** |
| Children: 1-5 years | 1640 [400-9,840] (40-126,040) | 440 [120-1,080] (40-15,880) | 1840 [240-19,940] (40-113,520) | 4120 [760-20,800] (40-80,360) |
| Children: 6-10 years | 760 [240-1,840] (40-61,560) | 320 [120-1,280] (40-73,360) | 520 [200-2,720] (40-48,600) | 1020 [320-5,100] (40-97,520) |
| Adolescents: 11-20 years | 320 [160-760] (40-27,440) | 180 [80-240] (40-42,120) | 280 [120-1,000] (40-42,880) | 440 [160-1,210] (40-66,760) |
| Adults: > 20 years | 200 [120-680] (40-31,040) | 120 [80-240] (40-41,320) | 120 [40-630] (40-40,280) | 220 [80-730] (40-19,040) |

Indoor residual spraying (IRS), Seasonal malaria chemoprevention (SMC)
[a] Number of participants surveyed that were analysed by microscopy.
[b] Data reflect the number (% (n/N)) of participants surveyed in each age group.
[c] Data reflect the number (% (n/N)) of participants surveyed that were microscopically positive for an asymptomatic *P. falciparum* infection (including mixed *P. falciparum* infections) relative to the number of participants surveyed in the total population and by the age groups presented.
[d] Median parasite density (i.e., parasites/µL of blood) [interquartile range (IQR)] (min-max) for the microscopically positive asymptomatic *P. falciparum* infections (including mixed *P. falciparum* infections).

**Table supplement 2.** The DBLα sequence pool sizes (upsA and non-upsA) and the number of unique DBLα types (upsA and non-upsA) observed in each survey (i.e., 2012, 2014, 2015, 2017) for those isolates with DBLα sequencing data.

| Survey | *P. falciparum* isolates sequenced | *P. falciparum* isolates with DBLα sequence data (≥ 1 DBLα type) [a] | *P. falciparum* isolates with DBLα sequence data (≥ 20 DBLα types) [a] | DBLα sequence pool size | | | Number of unique DBLα types | | |
|---|---|---|---|---|---|---|---|---|---|
| | | | | DBLα sequences | UpsA DBLα sequences [b] | Non-upsA DBLα sequences [c] | DBLα types | UpsA DBLα types [d] | Non-upsA DBLα types [e] |
| October 2012 (pre-IRS) | 808 | 742 (91.8) | 685 (84.8) | 120,029 | 22,881 (19.1) | 97,148 (80.9) | 35,377 | 2,218 (6.3) | 33,159 (93.7) |
| October 2014 (IRS) | 430 | 386 (89.8) | 301 (70.0) | 33,489 | 7,048 (21.0) | 26,441 (79.0) | 16,334 | 1,503 (9.2) | 14,831 (90.8) |
| October 2015 (post-IRS) | 545 | 510 (93.6) | 413 (75.8) | 42,774 | 8,942 (20.9) | 33,832 (79.1) | 19,584 | 1,673 (8.5) | 17,911 (91.5) |
| October 2017 (SMC) | 789 | 759 (96.2) | 700 (88.7) | 92,757 | 18,625 (20.1) | 74,132 (79.9) | 29,423 | 2,074 (7.0) | 27,349 (93.0) |
| TOTAL | 2,572 | 2,397 (93.2) | 2,099 (81.6) | 289,049 | 57,496 (19.9) | 231,553 (80.1) | 53,238 | 2,802 (5.3) | 50,436 (94.7) |

Indoor residual spraying (IRS), Seasonal malaria chemoprevention (SMC), Interquartile range (IQR)

[a] Data reflect the number (% (n/N)) of *P. falciparum* isolates that had DBLα sequencing data relative to the number of participants sampled that were positive for *P. falciparum* by microscopy (including mixed *P. falciparum* infections). For those a breakdown of those isolates by age group (years) with ≥ 20 DBLα types included in the analyses, see Table supplement 7.

[b] Data reflect the upsA DBLα sequence pool size (% (n/N)) relative to the DBLα sequence pool size.

[c] Data reflect the non-upsA DBLα sequence pool size (% (n/N)) relative to the DBLα sequence pool size.

[d] Data reflect the number (% (n/N)) of upsA DBLα types identified relative to the number of DBLα types identified.

[e] Data reflect the number (% (n/N)) of non-upsA DBLα types identified relative to the number of DBLα types identified.

**Table supplement 3.** The Kolmogorov-Smirnov Test (KS-test) is applied to compare the estimated MOI distributions for the population in each survey (i.e., 2012, 2014, 2015, 2017) for the two approaches, namely by pooling the maximum *a posteriori* MOI estimates or by using the mixture distributions (see Materials and Methods) for the uniform prior.

| Survey | Prior | KS-test statistics | *p-value* | Mean MOI difference [a] |
|---|---|---|---|---|
| October 2012 (pre-IRS) | Uniform | 0.019592 | 0.955175 | -0.13235 |
| October 2014 (IRS) | Uniform | 0.030432 | 0.943197 | -0.13112 |
| October 2015 (post-IRS) | Uniform | 0.038248 | 0.581476 | -0.1149 |
| October 2017 (SMC) | Uniform | 0.023823 | 0.821812 | -0.1298 |

Indoor residual spraying (IRS), Seasonal malaria chemoprevention (SMC)

[a] The mean MOI difference is the difference in the mean value of the population-level MOI estimates from either pooling the maximum *a posteriori* estimates or the mixture distribution.

**Table supplement 4.** The Kolmogorov-Smirnov Test (KS-test) is applied to compare the estimated MOI distributions for the population in each survey (i.e., 2012, 2014, 2015, 2017) for the two approaches, namely by pooling the maximum *a posteriori* MOI estimates or by using the mixture distributions (see Materials and Methods) using the negative binomial distribution with parameters in the range typical for low- (corresponding to a mean MOI ~ 1.5), medium- (corresponding to a mean MOI ~ 4.3), and high- (corresponding to a mean MOI ~ 6.7) transmission endemic areas.

| Survey | Negative binomial | KS-test statistics | *p-value* | Mean MOI difference [a] |
|---|---|---|---|---|
| October 2012 (pre-IRS) | Low | 0.024189 | 0.817681 | -0.14031 |
| | Medium | 0.021339 | 0.914045 | -0.11099 |
| | High | 0.019512 | 0.956709 | -0.11381 |
| October 2014 (IRS) | Low | 0.027862 | 0.973588 | -0.08387 |
| | Medium | 0.032505 | 0.908128 | -0.09739 |
| | High | 0.036612 | 0.814540 | -0.12282 |
| October 2015 (post-IRS) | Low | 0.037872 | 0.594217 | -0.09361 |
| | Medium | 0.032974 | 0.760228 | -0.11319 |
| | High | 0.037138 | 0.619209 | -0.11284 |
| October 2017 (SMC) | Low | 0.025665 | 0.745796 | -0.10952 |
| | Medium | 0.026964 | 0.68884 | -0.12406 |
| | High | 0.030275 | 0.542523 | -0.13645 |

Indoor residual spraying (IRS), Seasonal malaria chemoprevention (SMC)

[a] The mean MOI difference is the difference in the mean value of the population-level MOI estimates from either pooling the maximum *a posteriori* estimates or the mixture distribution for different parameter choices of a zero-truncated negative binomial prior.

**Table supplement 5.** The Kolmogorov-Smirnov Test (KS-test) and the Pearson correlation tests (PC-test) are applied to compare the estimated MOI distributions for the population in each survey (i.e., 2012, 2014, 2015, 2017) based on either a uniform prior or a zero-truncated negative binomial prior with parameters in the ranges typical for low- (corresponding to a mean MOI ~ 1.5), medium- (corresponding to a mean MOI ~ 4.3), and high- (corresponding to a mean MOI ~ 6.7) transmission endemic areas (see Materials and Methods). We present the results of comparison for both approaches which obtain the population-level MOI distribution from individual posterior MOI distributions, namely by pooling the maximum *a posteriori* MOI estimates (i.e., MAP pool) or by using mixture distribution (i.e., Mixture Dist.).

| Survey | Approach | Comparison (Negative binomial vs. Uniform) | KS-test statistics | KS-test *p-value* | PC-test statistics | PC-test *p-value* | Mean MOI difference [a] |
|---|---|---|---|---|---|---|---|
| October 2012 (pre-IRS) | MAP pool | Low vs. Uniform | 0.054015 | 0.036738 | 0.988473 | 0 | -0.32117 |
| | MAP pool | Medium vs. Uniform | 0.018978 | 0.966014 | 0.994902 | 0 | -0.08029 |
| | MAP pool | High vs. Uniform | 0.018978 | 0.966014 | 0.997470 | 0 | -0.00438 |
| October 2014 (IRS) | MAP pool | Low vs. Uniform | 0.036545 | 0.816283 | 0.990630 | 1.4E-260 | -0.13289 |
| | MAP pool | Medium vs. Uniform | 0.016611 | 0.999997 | 0.996893 | 0 | -0.00332 |
| | MAP pool | High vs. Uniform | 0.019934 | 0.999760 | 0.997409 | 0 | 0.01661 |
| October 2015 (post-IRS) | MAP pool | Low vs. Uniform | 0.046005 | 0.346351 | 0.984101 | 0 | -0.15496 |
| | MAP pool | Medium vs. Uniform | 0.009685 | 1 | 0.99500 | 0 | -0.02179 |
| | MAP pool | High vs. Uniform | 0.024213 | 0.968793 | 0.996081 | 0 | 0.01937 |
| October 2017 (SMC) | MAP pool | Low vs. Uniform | 0.055714 | 0.025924 | 0.985225 | 0 | -0.22714 |
| | MAP pool | Medium vs. Uniform | 0.011429 | 0.999989 | 0.994265 | 0 | -0.04714 |
| | MAP pool | High vs. Uniform | 0.011429 | 0.999989 | 0.996798 | 0 | -0.00286 |
| October 2012 (pre-IRS) | Mixture Dist. | Low vs. Uniform | 0.05387 | 0.037968 | - | - | -0.31321 |
| | Mixture Dist. | Medium vs. Uniform | 0.015199 | 0.997437 | - | - | -0.10165 |
| | Mixture Dist. | High vs. Uniform | 0.007393 | 1 | - | - | -0.02292 |
| October 2014 (IRS) | Mixture Dist. | Low vs. Uniform | 0.040398 | 0.709761 | - | - | -0.18014 |
| | Mixture Dist. | Medium vs. Uniform | 0.011440 | 1 | - | - | -0.03705 |
| | Mixture Dist. | High vs. Uniform | 0.010149 | 1 | - | - | 0.00832 |
| October 2015 (post-IRS) | Mixture Dist. | Low vs. Uniform | 0.049573 | 0.263382 | - | - | -0.17625 |
| | Mixture Dist. | Medium vs. Uniform | 0.008205 | 1 | - | - | -0.02350 |
| | Mixture Dist. | High vs. Uniform | 0.010618 | 1 | - | - | 0.01732 |
| October 2017 (SMC) | Mixture Dist. | Low vs. Uniform | 0.048020 | 0.07961 | - | - | -0.24742 |
| | Mixture Dist. | Medium vs. Uniform | 0.009626 | 1 | - | - | -0.05289 |
| | Mixture Dist. | High vs. Uniform | 0.009381 | 1 | - | - | 0.00379 |

Indoor residual spraying (IRS), Seasonal malaria chemoprevention (SMC), NA (-)

[a] The mean MOI difference is the difference in the mean value of the population-level MOI estimates from assuming either a zero-truncated negative binomial prior with different parameter choices, or a uniform prior. Take the first row of the table as an example, the mean MOI difference value (-0.32117) is equal to the mean MOI for the population based on a zero-truncated negative binomial prior with a low-transmission parameter choice minus that based on a uniform prior. The order of comparison is consistent with the one listed in the Comparison column (Low vs. Uniform).

**Table supplement 6.** The Kolmogorov-Smirnov Test (KS-test) and the Pearson correlation tests (PC-test) are applied to compare the estimated MOI distributions for the population in each survey (i.e., 2012, 2014, 2015, 2017) between the zero-truncated negative binomial priors with parameters in the ranges typical for low- (corresponding to a mean MOI ~ 1.5), medium- (corresponding to a mean MOI ~ 4.3), and high- (corresponding to a mean MOI ~ 6.7) transmission endemic areas (see Materials and Methods). We present the results of comparison for both approaches which obtain the population-level MOI distribution from individual posterior MOI distributions, namely by pooling the maximum *a posteriori* MOI estimates (i.e., MAP pool) or by using mixture distribution (i.e., Mixture Dist.).

| Survey | Approach | Comparison (Negative binomial) | KS-test statistics | KS-test *p-value* | PC-test statistics | PC-test *p-value* | Mean MOI difference [a] |
|---|---|---|---|---|---|---|---|
| October 2012 (pre-IRS) | MAP pool | Medium vs. Low | 0.051095 | 0.055938 | 0.986680 | 0 | 0.240876 |
| | MAP pool | Medium vs. High | 0.014599 | 0.998598 | 0.996125 | 0 | -0.075912 |
| | MAP pool | Low vs. High | 0.055474 | 0.029513 | 0.986516 | 0 | -0.316788 |
| October 2014 (IRS) | MAP pool | Medium vs. Low | 0.053156 | 0.362791 | 0.989269 | 8.011E-252 | 0.129568 |
| | MAP pool | Medium vs. High | 0.003322 | 1 | 0.998276 | 0 | -0.019934 |
| | MAP pool | Low vs. High | 0.056478 | 0.292237 | 0.988379 | 1.112E-246 | -0.149502 |
| October 2015 (post-IRS) | MAP pool | Medium vs. Low | 0.055690 | 0.154269 | 0.981405 | 9.764E-297 | 0.133172 |
| | MAP pool | Medium vs. High | 0.014528 | 0.999994 | 0.994613 | 0 | -0.041162 |
| | MAP pool | Low vs. High | 0.070218 | 0.034065 | 0.979963 | 3.920E-290 | -0.174334 |
| October 2017 (SMC) | MAP pool | Medium vs. Low | 0.044286 | 0.128371 | 0.983760 | 0 | 0.18 |
| | MAP pool | Medium vs. High | 0.011429 | 0.999989 | 0.995738 | 0 | -0.044286 |
| | MAP pool | Low vs. High | 0.055714 | 0.025924 | 0.982745 | 0 | -0.224286 |
| October 2012 (pre-IRS) | Mixture Dist. | Medium vs. Low | 0.040399 | 0.214223 | - | - | 0.211562 |
| | Mixture Dist. | Medium vs. High | 0.011531 | 0.999989 | - | - | -0.078734 |
| | Mixture Dist. | Low vs. High | 0.052049 | 0.049412 | - | - | -0.290295 |
| October 2014 (IRS) | Mixture Dist. | Medium vs. Low | 0.037816 | 0.785935 | - | - | 0.143090 |
| | Mixture Dist. | Medium vs. High | 0.011447 | 1 | - | - | -0.045364 |
| | Mixture Dist. | Low vs. High | 0.050535 | 0.425625 | - | - | -0.188454 |
| October 2015 (post-IRS) | Mixture Dist. | Medium vs. Low | 0.047779 | 0.303800 | - | - | 0.152752 |
| | Mixture Dist. | Medium vs. High | 0.010639 | 1 | - | - | -0.040816 |
| | Mixture Dist. | Low vs. High | 0.061610 | 0.087626 | - | - | -0.193568 |
| October 2017 (SMC) | Mixture Dist. | Medium vs. Low | 0.042425 | 0.161444 | - | - | 0.194537 |
| | Mixture Dist. | Medium vs. High | 0.0096285 | 0.999999 | - | - | -0.056674 |
| | Mixture Dist. | Low vs. High | 0.052140 | 0.044719 | - | - | -0.251211 |

Indoor residual spraying (IRS), Seasonal malaria chemoprevention (SMC), NA (-)

[a] The mean MOI difference is the difference in the mean value of the population-level MOI estimates from assuming zero-truncated negative binomial priors with different parameter choices. Take the first row of the table as an example, the mean MOI difference value (0.240876) is equal to the mean MOI for the population based on a zero-truncated negative binomial prior with a medium-transmission parameter choice minus that based on a zero-truncated negative binomial prior with a low-transmission parameter choice. The order of comparison is consistent with the one listed in the Comparison column (Medium vs. Low).

**Table supplement 7.** Microscopic *P. falciparum* DBLα type sequencing results, number of *P. falciparum var* repertoires (i.e., census population size), and mean $MOI_{var}$ by age group (years) in each survey (i.e., 2012, 2014, 2015, 2017).

| | October 2012 (pre-IRS) | October 2014 (IRS) | October 2015 (post-IRS) | October 2017 (SMC) |
|---|---|---|---|---|
| Number of microscopic *P. falciparum* isolates | **808** | **430** | **545** | **789** |
| *P. falciparum* isolates with DBLα sequencing data (≥ 20 DBLα types) [a*] | **685 (84.8)** | **301 (70.0)** | **413 (75.8)** | **700 (88.7)** |
| Children: 1-5 years | 158 (91.3) | 28 (75.7) | 51 (81.0) | 44 (89.8) |
| Children: 6-10 years | 217 (89.3) | 116 (81.7) | 146 (87.4) | 170 (92.4) |
| Adolescents: 11-20 years | 167 (82.7) | 112 (69.1) | 129 (76.3) | 284 (93.4) |
| Adults: >20 years | 143 (75.3) | 45 (50.6) | 87 (59.6) | 202 (68.5) |
| Number of *P. falciparum var* repertoires (i.e., census population size) [b*] | **2552 (2354 - 2756)** | **731 (628 - 836)** | **909 (804 - 1019)** | **2087 (1926 - 2254)** |
| Children: 1-5 years | 495 (402 - 595) | 80 (43-125) | 62 (39 - 88) | 90 (54 - 132) |
| Children: 6-10 years | 1035 (908 - 1166) | 318 (247 - 397) | 378 (311 - 451) | 750 (642 - 861) |
| Adolescents: 11-20 years | 683 (579 - 792) | 256 (200 - 315) | 313 (250 - 382) | 827 (734 - 921) |
| Adults: >20 years | 338 (278 - 401) | 76 (48 - 111) | 155 (119 - 195) | 420 (365 - 478) |
| *P. falciparum* mean $MOI_{var}$ [c*] | **4.38 (4.16 - 4.61)** | **2.74 (2.49 - 3.02)** | **2.57 (2.40 - 2.77)** | **3.28 (3.12 - 4.45)** |
| Children: 1-5 years | 5.16 (4.68 - 5.67) | 2.86 (1.93 - 4.04) | 2.27 (1.98 - 2.59) | 3.34 (2.73 - 4.05) |
| Children: 6-10 years | 5.26 (4.87 - 5.67) | 3.22 (2.76 - 3.72) | 2.96 (2.64 - 3.32) | 4.41 (4.00 - 4.84) |
| Adolescents: 11-20 years | 4.09 (3.68 - 4.52) | 2.59 (2.25 - 2.96) | 2.74 (2.40 - 3.12) | 3.45 (3.21 - 3.70) |
| Adults: >20 years | 2.52 (2.27 - 2.78) | 1.80 (1.38 -2.38) | 1.85 (1.62 - 2.11) | 2.08 (1.93 - 2.24) |

[a] Data reflect the number (% (n/N)) of *P. falciparum* isolates that had DBLα sequencing data relative to the number of participants sampled that were positive for *P. falciparum* by microscopy (including mixed *P. falciparum* infections) in the total population and by the age groups (see Table 1 for total number of participants sampled that were microscopy positive).

[b] Number of *var* repertoires (i.e., census population size) (95% confidence interval). To account for differences in sampling depth across age groups and surveys, we performed subsampling with replacement by selecting the minimum number of individuals in each age group across all surveys. We then calculated the total number of *var* repertoires from these subsampled individuals within each age group in each survey. This approach ensures consistent sample sizes within each age group across all surveys. Finally, we summed the *var* repertoires across age groups to obtain the total *var* repertoire count for each survey. The mean and 95% CIs for the number of *var* repertoires (i.e., census population size) were estimated by repeating the subsampling procedure 10,000 times. The CIs were then derived from the distribution of these repeated subsampling replicates.

[c] Mean $MOI_{var}$ (95% confidence interval) based on pooling the maximum *a posteriori* MOI estimates. The 95% confidence intervals (95% CIs) were calculated based on a bootstrap approach. We resampled 10,000 replicates from the original population-level MOI distribution with replacement. Each resampled replicate has the same size as the original sample. We then derive the 95% CI based on the distribution of the resampled replicates.

[*] *Note: There is a simple map between census population size and mean MOI, as one can simply divide or multiply by the sample size (i.e., the number of P. falciparum isolates with DBLα sequencing data), respectively, to convert between the two quantities.*
